# Cross-System Legibility of a Practitioner-Derived Workflow-Error Taxonomy for Conversational AI: A Three-Comparator Agreement Study

**DOI:** 10.64898/2026.09.02.26362090

**Authors:** Davis Austria, Byrron McCollister, Jeremy E. Lindsey, Micheal Arowolo, Marian Okon

## Abstract

Practitioner-derived taxonomies of conversational artificial intelligence (AI) workflow errors show low inter-rater agreement among human coders, leaving open whether the instrument is ill-specified or the judgments are inherently difficult. We delivered a locked eight-category workflow-error taxonomy verbatim, under standardized conditions, to three frontier large language model comparators from distinct developer lineages, which coded a documented 45-incident error corpus. On the same 16 incidents coded by three human reviewer-authors, comparator category agreement was substantial (Fleiss κ=0.625) against slight human agreement (κ=0.155); across the full corpus it was stable (κ=0.632), and a 10-category refinement did not reduce it (κ=0.690). Severity and a claimed-verification flag remained only fair in both arms and on both samples. Substantial cross-system consistency provides a legibility signal consistent with recoverable category distinctions, but cannot separate instrument clarity from shared model priors, and is not a validation of any coding.

## Introduction

Benchmark evaluations of large language models (LLMs) score isolated outputs, while deployed failures occur inside multi-step workflows where erroneous content propagates into documents, analyses, protocols, and decisions ^1^. Documented failure modes of artificial intelligence (AI) systems such as hallucination^2^ and sycophancy^3^ are typically measured at the prompt level, using curated items with known answers; recent work argues that evaluating interactive systems requires designs beyond static benchmarks and end-to-end audit frameworks^4,5^, and the instruments available for characterizing errors as they occur inside real research and clinical informatics work — with downstream consequence, recurrence, and correction burden — are earlier in their development. This mirrors a familiar arc in patient safety measurement, where incident taxonomies and structured error models preceded, and eventually enabled, reliable surveillance^6,7^: the usefulness of any error taxonomy depends first on whether independent coders can apply it consistently.

A practitioner-audit framework (TRACE: Tracking Reliability of AI-generated Conversational Evidence) recently proposed an eight-category workflow-error taxonomy with a consequence-based severity rubric and a claimed-verification flag, demonstrated on a 45-incident documented error corpus recorded in the reflexive-autoethnographic tradition^1^. In that demonstration, independent category agreement among three human reviewer-authors on a stratified 16-incident subsample was slight (Fleiss κ=0.155)^8^, with the claimed-verification flag near chance (κ=0.005). The reviewers converged on a diagnosis: overlapping categories, missing codes, and an under-specified claimed-verification dimension.

Low human agreement admits two readings. The instrument may be ill-specified — its category boundaries not recoverable from its text — or the judgments may be inherently difficult even for a well-specified instrument, because human coders bring divergent priors about severity, consequence, and intent, and read incidents against different professional histories. The two readings carry different remedies: the first calls for instrument revision; the second for coder training, written adjudication conventions, or acceptance that the construct carries an irreducible judgment component. Distinguishing them requires a measurement design in which coder-side variance is minimized while the instrument is held fixed.

Independently developed AI systems offer such a probe. When structurally distinct LLMs from different developer lineages receive an identical instrument under standardized delivery conditions — identical system prompt, fixed decoding temperature, structured output schema — their cross-system agreement provides a signal about how far the instrument’s distinctions are recoverable from its text alone, stripped of the individual histories human coders bring. We term this signal taxonomy legibility. It is explicitly not validation: agreement among AI systems cannot establish that any code is correct, and systems sharing training corpora may agree for reasons unrelated to instrument clarity^9,10^. Used with those limits stated, however, legibility probing addresses a question human panels cannot cleanly answer about their own disagreement, at low marginal cost, under an auditable and standardized replication protocol.

We asked three questions. (1) How consistently do three LLM comparators from distinct developer lineages apply the locked eight-category taxonomy across the full 45-incident corpus? (2) Do severity and the claimed-verification flag — the judgment-heavy dimensions — show the same pattern as category assignment, or dissociate from it? (3) Does category agreement survive a reviewer-motivated refinement of the category set from eight to ten codes?

## Methods

Corpus. The coding objects were the 45 documented error incidents of the TRACE demonstration corpus: errors involving one frontier conversational LLM, recorded by a clinician-informatician practitioner over seven weeks (March 15 to May 7, 2026) across scholarly, clinical informatics, and clinical-adjacent research workflows, including evidence synthesis screening, manuscript development, credentialing and conference materials, regulatory and institutional review board drafting, and public-health program review^1^. Each incident carries a de-identified error summary, the correction source, and the documented downstream consequence; incidents entered the corpus under documentation-positive criteria (consequential enough to detect and record), so the corpus is harm-weighted rather than prevalence-weighted. The audited conversational system was excluded from the comparator set to avoid self-assessment circularity, and comparator systems are de-identified consistent with the companion work’s vendor-blinding rationale: the object of study is the instrument, not any vendor.

Instrument. The locked eight-category taxonomy was delivered with one-line operational anchors (Table 1), together with a four-level consequence-based severity rubric (Critical, High, Medium, Low, coded from downstream workflow consequence rather than output form) and a three-value claimed-verification flag (Yes, No, Unclear) capturing whether the erroneous output claimed or implied that a verification act had occurred. Each comparator was instructed to act as a blinded independent coder, to code each incident independently without revising earlier codings, and to return exactly one JavaScript Object Notation (JSON) object per incident containing the incident identifier, category, severity, claimed-verification value, a descriptive 0.0–1.0 confidence (stated to be non-calibrated), and a brief methodological note.

**Table 1.** Eight-category instrument delivered to comparators (operational anchors).

| Category | Operational anchor |
| --- | --- |
| Verification failure | False or implied performance of a verification process: the output claims or implies a check occurred, or asserts direct access to an artifact's state, when no adequate check took place |
| Factual numerical error | Wrong number, date, value, or fabricated quantitative content |
| Tool-behavior misunderstanding | Misrepresentation of platform, tool, format, or system behavior or capability |
| Unverified claim presented as fact | Unsupported factual assertion without an explicit or implied claim that verification occurred |
| Citation or reference formatting error | Citation, reference, terminology, or regulatory-citation defect, including reference-apparatus omissions |
| Workflow contradiction | Contradictory or shifting procedural guidance, including loss of prior task constraints |
| Document or protocol-safety failure | Structural document-integrity damage or unsafe protocol guidance |
| Identity or positionality | Misassigned identity, attribution, or identifier |

Comparators and runs. Three structurally distinct frontier LLM comparators from three different developer lineages were accessed via hosted application programming interfaces (APIs) at decoding temperature 0; where a system exposed deterministic seeding it was fixed, and otherwise outputs retain run-to-run stochasticity. Comparators are reported as Systems A, B, and C. Identities are withheld by design: the audited system of the companion work was excluded from the comparator set, so naming the comparators would narrow the audited system by elimination and compromise that study’s vendor blinding. Full run parameters — verbatim system prompt, output schema, temperature, token limits, normalization rules, and run dates — are reported so that the protocol can be replicated against any three frontier systems; exact identifiers are available to editors under confidentiality. Coding proceeded in two rounds: Round A coded a 16-incident stratified random subsample (stratified by severity; seed 20260509) matching the human-coded subsample exactly, and Round B coded the remaining 29 incidents under identical conditions. Round A provides the matched human-versus-comparator contrast; the pooled 45-incident analysis provides the full-corpus result.

Analysis. Category agreement across the three comparators used Fleiss κ^8^, with pairwise Cohen κ^11^ for each comparator pair; category labels were compared case-insensitively after stripping any leading enumeration prefix. Severity used Fleiss κ over the four ordered levels and the claimed-verification flag used Fleiss κ over its three values. The companion work reported human severity agreement pairwise with quadratic-weighted κ rather than as a three-rater Fleiss value; the two statistics are not directly comparable, and human severity is therefore not contrasted against the comparator severity statistic here. Agreement magnitudes are described using the Landis and Koch benchmarks^12^, acknowledging their conventional rather than principled thresholds. The primary human-versus-comparator contrast is computed on the identical 16 incidents coded by both arms; the 45-incident pooled analysis is reported as a full-corpus extension rather than as a direct contrast against the human statistic. Human coding of record is the three reviewer-authors’ independent coding of that subsample, each blinded to the practitioner-author’s coding and to one another, reported in the companion work^1^. A standalone sensitivity analysis re-ran all 45 incidents under a 10-category instrument adding two reviewer-motivated categories (identifier error; behavioral or interactional pattern); because the 8- and 10-category runs are separate and unpaired, the sensitivity result is interpreted only as whether refinement reduced legibility, not as a demonstrated improvement.

Reflexivity, AI assistance, and ethics. Consistent with the companion work’s reflexive tradition, the analysis pipeline itself was AI-assisted under human verification: comparator API runners, parsing, and agreement computations were drafted with an AI editing assistant and verified by the practitioner-author against hand-checked cases, with all scripts, prompt logs, parsed codings, and kappa reports retained in the study audit trail. The corpus consists of the practitioner-author’s own documented workflow incidents, de-identified; no patient data and no human subjects beyond the authors are involved. The comparator analysis was framed to the comparators, and is reported here, strictly as an interpretability check whose outputs assess the taxonomy rather than revise the human coding of record.

## Results

Matched-sample comparison. On the 16 incidents coded by both arms, the three comparators agreed substantially on category assignment (Fleiss κ=0.625), against slight agreement among the three human reviewer-authors on the same incidents (Fleiss κ=0.155). Pairwise comparator category agreement on this sample ranged from κ=0.564 to κ=0.769. The dimension-specific pattern was already present here: severity agreement was fair (κ=0.253) and claimed-verification agreement fair (κ=0.387), well below category agreement in the same systems on the same incidents (Figure 1).

**Figure 1.**
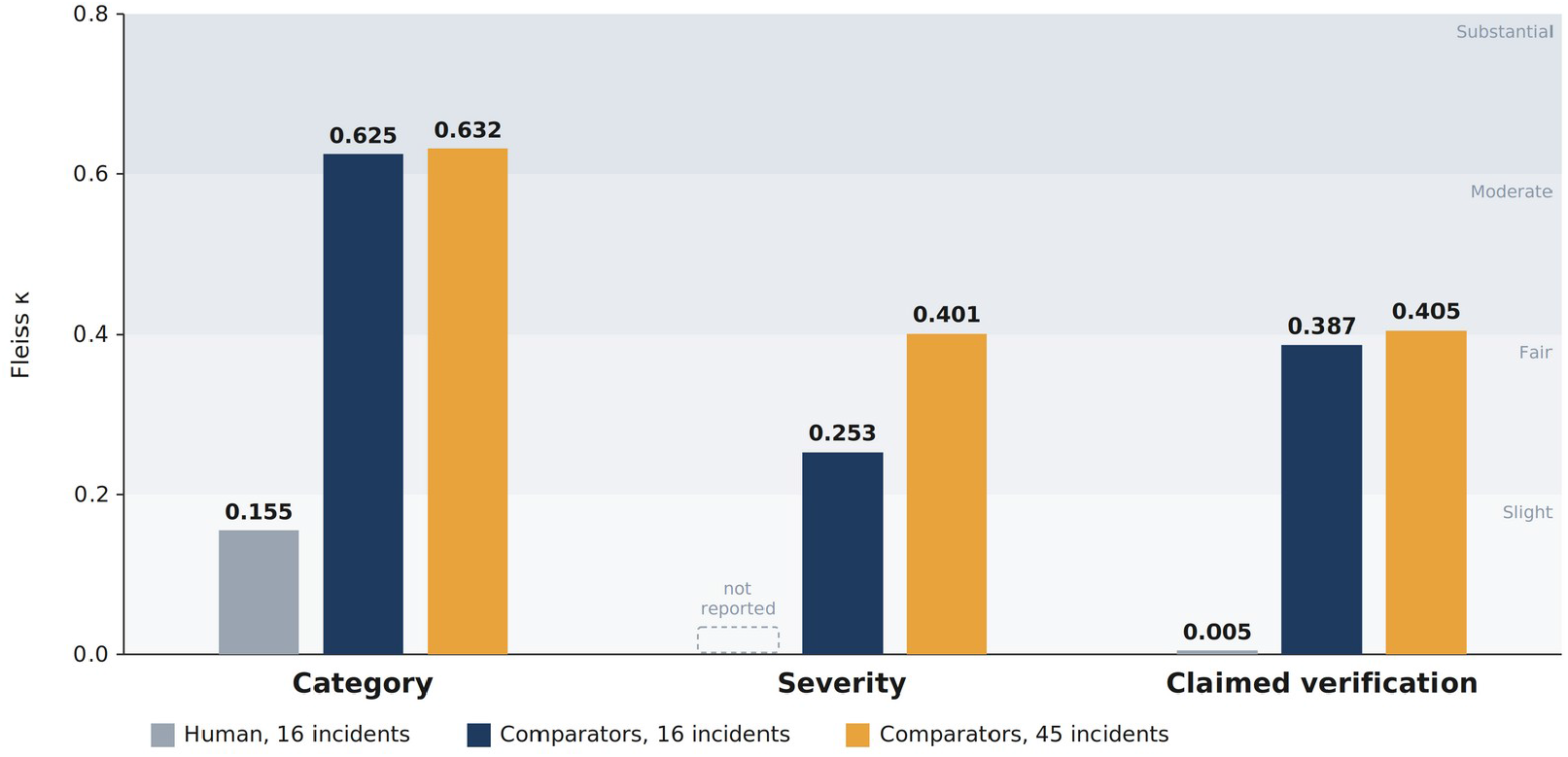
Dimension-specific agreement across arms and samples. On the 16 incidents coded by both arms, comparator category agreement (Fleiss κ=0.625) exceeded human agreement (κ=0.155); the comparator value was stable across the full 45-incident corpus (κ=0.632). In both arms and on both samples, severity and claimed verification were applied less consistently than category. Human severity agreement was computed pairwise with quadratic-weighted kappa (0.034–0.636), which credits nearmiss ratings on an ordinal scale and is not directly comparable to the nominal Fleiss statistic used for the comparator arm; no three-rater Fleiss value for human severity was computed. Shaded bands mark Landis and Koch benchmarks.

**Table 2.**
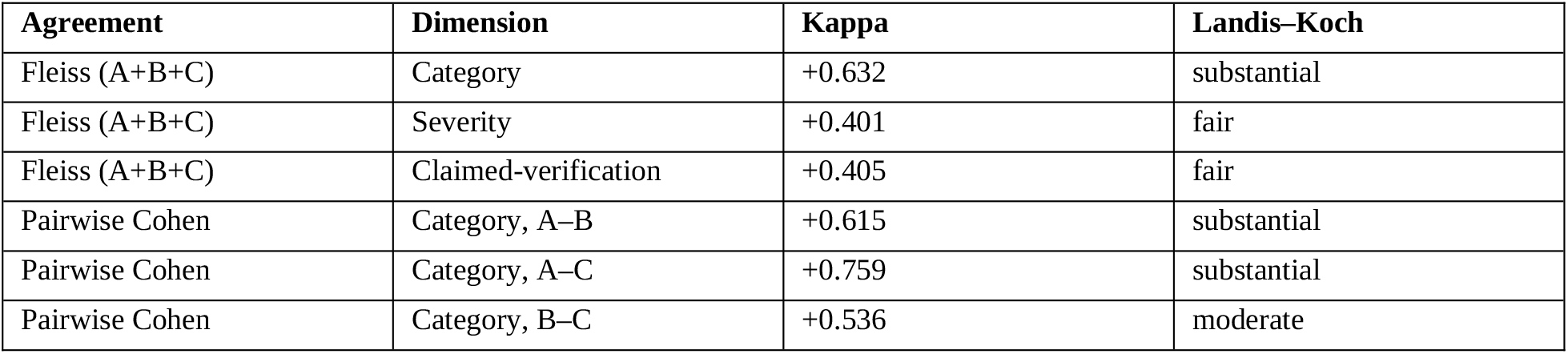
Cross-comparator agreement, eight-category instrument, 45 incidents.

Full-corpus extension. Across all 45 incidents under the eight-category instrument, comparator category agreement was stable at Fleiss κ=0.632, with pairwise Cohen κ of 0.615 (A–B), 0.759 (A–C), and 0.536 (B–C); severity agreement was fair (κ=0.401), as was the claimed-verification flag (κ=0.405). The close correspondence between the matched-sample and full-corpus category values (0.625 and 0.632) suggests that the comparator result was not driven by the subsample composition. Human pairwise category agreement on the matched incidents ranged from κ=0.055 to κ=0.566, indicating that the low panel statistic was not driven by a single outlying coder. In both arms, and on both samples, category agreement exceeded agreement on severity and claimed verification.

Sensitivity analysis. Under the 10-category instrument, comparator category agreement did not fall and was somewhat higher (Fleiss κ=0.690); severity (κ=0.364) and claimed-verification (κ=0.363) remained fair. All three comparators assigned the two added categories predominantly to the incidents that had motivated the reviewers to propose them. Because the runs are unpaired, this is read as refinement not reducing category-level legibility, rather than as an improvement.

## Discussion

Principal finding. On the same 16 incidents, three structurally distinct systems given nothing but the locked definitions under standardized delivery classified incidents with substantial mutual consistency (κ=0.625), where three human reviewer-authors agreed only slightly (κ=0.155); the comparator value held across the full corpus (κ=0.632) and was not reduced by a reviewer-motivated refinement of the category set. The result constrains one explanation of the low human agreement observed in the companion demonstration^1^: it cannot be attributed straightforwardly to the category distinctions being unrecoverable from the instrument’s text, since independently developed systems reading only that text produced substantially consistent classifications. It does not, however, decompose instrument-side from judgment-side variance, because consistent machine classification may also arise from shared model priors. What the data show more directly is a dimension-specific dissociation: in both arms and on both samples, category assignment was more consistently applied than severity or claimed verification, consistent with those dimensions carrying a judgment component that operational definitions have not yet captured.

Relation to prior work. A growing literature reports that LLMs can match or exceed human annotators on text-classification tasks^9,10^. The present design differs in aim: rather than asking whether AI coders can replace human ones, it uses cross-system agreement as an instrument-development diagnostic — a diagnostic signal for investigating whether low human reliability may reflect instrument-side or judgment-side difficulty. That use inherits the literature’s caveats (shared priors, prompt sensitivity) but avoids its strongest claim, since no coding here is treated as correct.

Design rationale for legibility probes. Several design choices generalize to other instrument developers. Delivering the instrument verbatim in the system prompt, rather than paraphrasing it, keeps the probe about the instrument’s actual text. Decoding at temperature 0, with seeds fixed where available, minimizes within-system variance so that residual disagreement is attributable to the systems’ readings rather than sampling. Requiring structured JSON output with a forced single category prevents hedged multi-label responses that would inflate apparent agreement ambiguity. Drawing comparators from distinct developer lineages weakens, though does not eliminate, the shared-prior confound. Excluding the audited system from the comparator set removes self-assessment circularity. Finally, running a stratified subsample first (Round A) before committing the full corpus (Round B) allows an inexpensive early check that the output schema, normalization, and agreement pipeline behave as intended before the primary analysis is generated.

Cautions. Three cautions govern interpretation. First, agreement is not accuracy: cross-comparator consistency cannot establish that any coding is right, and no ground truth exists for these constructs beyond adjudicated practitioner coding. Second, the comparators may share overlapping training corpora and architectural ancestry; their agreement may partly reflect shared model priors rather than instrument clarity, which is why the analysis is framed as triangulation against, not confirmation of, the human coding. Third, the human coders worked from richer context, and their disagreements may encode real ambiguity in the incidents that standardized delivery suppresses; suppressing ambiguity is not resolving it, and a legibility probe cannot distinguish the two.

Implications. For informatics evaluation practice, the method offers a low-cost, auditable legibility probe for error-taxonomy development: when a new instrument shows low human agreement, delivering it to AI comparators from distinct developer lineages indicates whether the category distinctions are at least recoverable from the instrument text, before investing in coder training or instrument revision. For the TRACE program specifically, the results support retaining the eight-category structure, directing definitional work at the severity and claimed-verification dimensions — whose sharpened operational conventions are now documented in the companion work — and testing in a designed replication whether those conventions improve human agreement toward levels observed under standardized comparator application.

Limitations. The corpus derives from a single practitioner and a single audited system, and its documentation-positive construction weights consequential incidents. The comparator set is small, may not span current model diversity, and may share training corpora or architectural ancestry, so cross-system consistency cannot be read as independent confirmation. Prompt-format sensitivity was not tested; where deterministic seeding was unavailable, outputs retain run-to-run stochasticity, and hosted models may change over time, so replication is standardized rather than exact. The matched contrast rests on 16 incidents, and the 8-versus 10-category comparison is unpaired. Finally, legibility probing as proposed here is itself a candidate method requiring evaluation across instruments and domains.

## Conclusion

A practitioner-derived workflow-error taxonomy for conversational AI was applied with substantial consistency by three LLM comparators from distinct developer lineages under standardized conditions, on the same incidents where human reviewer-author agreement was slight (κ=0.625 versus κ=0.155; κ=0.632 across the full corpus, sustained at κ=0.690 under a 10-category refinement). Its judgment-heavy dimensions — severity and claimed verification — were not, in either arm or on either sample. The dissociation supports prioritizing definitional refinement of the judgment-heavy dimensions before further category redesign, and offers AI-comparator legibility probing as a reusable, low-cost diagnostic for error-taxonomy development in biomedical informatics, with the caveat that cross-system consistency cannot by itself separate instrument clarity from shared model priors.

## Data Availability

The comparator instrument, response schema, normalization rules, and all agreement statistics reported here are contained in the manuscript and in the companion work's supplementary materials (doi:10.64898/2026.08.13.26360414). De-identified per-incident comparator codings, analysis scripts, prompt logs, and kappa reports are retained in the study audit trail and are available upon reasonable request to the corresponding author. Exact comparator model identifiers are withheld from the published record to preserve the vendor blinding of the companion study and are available to editors under confidentiality; the full run protocol is reported so that the design can be replicated against any three independently developed frontier systems.

https://www.medrxiv.org/content/10.64898/2026.08.13.26360414v1

## Data and materials availability

The verbatim comparator instrument, response schema, normalization rules, and the agreement statistics reported here are provided in the companion work’s supplementary materials^1^. De-identified per-incident comparator codings, analysis scripts, prompt logs, and kappa reports are retained in the study audit trail and are available from the corresponding author for editorial review and reproducibility checking. Exact comparator identifiers are withheld from the published record for the reason stated in the Methods and are available to editors under confidentiality; the full run protocol is reported so that the design can be replicated against any three independently developed frontier systems.

